# Individuals with Parkinson’s Disease Retain Spatiotemporal Gait Control With Music and Metronome Cues

**DOI:** 10.1101/19003269

**Authors:** Guneet Chawla, Madelon Wygand, Nina Browner, Michael D Lewek

## Abstract

**Background:** Parkinson’s disease (PD) is marked by a loss of motor automaticity, resulting in decreased control of step length during gait. Rhythmic auditory cues (metronomes or music) may enhance automaticity by adjusting cadence. Both metronomes and music may offer distinct advantages, but prior attempts at quantifying their influence on spatiotemporal aspects of gait have been confounded by altered gait speeds from overground walking. We hypothesized that when gait speed is fixed, individuals with PD would experience difficulty in modifying cadence due to the concomitant requirement to alter step length, with greater changes noted with metronomes compared to music cues.

**Research Question:** Can a metronome or music promote spatiotemporal adjustments when decoupled from changes in gait speed in individuals with PD?

**Methods:** 21 participants with PD were instructed to time their steps to a metronome and music cues (at 85%, 100%, and 115% of overground cadence) during treadmill walking. We calculated cadence, cadence accuracy, and step length during each cue condition and an uncued control condition. We compared the various cue frequencies and auditory modalities.

**Results:** At fixed gait speeds, participants were able to increase and decrease cadence in response to auditory cues. Music and metronome cues produced comparable results in cadence manipulation with greater cadence errors noted at slower intended frequencies. Nevertheless, the induced cadence changes created a concomitant alteration in step length, with music and metronomes producing comparable changes. Notably, longer step lengths were induced with both music and metronome during slow frequency cueing.

**Significance:** This important change conflicts with conventional prescriptive approaches, which advocate for faster cue frequencies, if applied on a treadmill. The music and metronome cues produced comparable changes to gait, suggesting that either cue may be effective at overcoming the shortened step lengths during treadmill walking if slower frequencies are used.

## Introduction

The characteristic short, shuffling steps and tendency of individuals with Parkinson’s disease (PD) to freeze during walking are often attributed to diminished automaticity [1-3]. Rhythmic auditory cueing is a well-established intervention intended to address these deficits in gait automaticity by adjusting cadence [4]. Previous studies have used either music or a metronome to provide the auditory cues [2, 5]. Despite the potential advantages to both metronomes and music, prior attempts at quantifying their influence on spatiotemporal aspects of gait have been confounded by altered gait speeds from overground walking [6]. Thus, it remains unclear if either auditory modality can promote the intended spatiotemporal adjustments when decoupled from changes in gait speed.

A key benefit of a metronome is that the beat is discrete, enabling synchronization between the individual’s steps and the beat [5, 7]. Music, however, which may be more enjoyable than a metronome and is used commonly during exercise, may be less effective than the metronome due to the many layers of rhythms and beats that make musical sounds continuous rather than discrete [8]. Any difficulty in identifying the beat of a song may make it more challenging to adjust step timing appropriately. Nevertheless, several studies have examined the effect of music on gait mechanics, and have found a generally favorable association between the use of music cues and increased stride length [2, 9-11]. Potentially confounding these promising results, however, is the fact that these studies typically used a single musical track that was engineered to accentuate the rhythm to enhance detection. Nevertheless, direct comparisons of gait mechanics while stepping to either music or metronome cues are rare. In studies involving healthy older adults, metronome cues elicit greater synchronization compared to music cues [7]. However, both music and metronome cues effectively increased cadence, with only the musical cues leading to a concomitant increase in stride length and gait velocity of healthy older adults [12].

Although the use of auditory cues has the potential to improve gait in individuals with PD, we are unaware of evidence to support the preferential use of music or metronomes in this population [5]. The purpose of this study was therefore to determine the difference in spatiotemporal gait measures induced by stepping to the beat of a metronome and to music cues of various frequencies. Importantly, prior work examining the use of auditory cues has involved over-ground walking, [10] allowing participants to change gait speed in response to different frequency cues. Because cadence and step length are both influenced by gait speed, [13] we used treadmill gait to carefully assess potential changes in spatiotemporal gait parameters across various auditory cue conditions, without the confounding influence of altered gait speed. Given the shortened step lengths observed in individuals with PD [14, 15], we were particularly concerned that individuals would exhibit difficulty in decreasing their cadence on the treadmill, because this would require a necessary increase in step length. Because gait speed was fixed on the treadmill, we hypothesized that attempts to decrease cadence (and increase step length) would be more challenging than attempts to increase cadence (and decrease step length). We further hypothesized that an inability to discern the tempo in the music accurately would reduce cadence accuracy compared to stepping with a metronome [7]. As a result, we believe that changes in spatiotemporal measures will be particularly evident with metronome cues compared to music cues.

## Methodology

### Procedures

We recruited individuals from the UNC Movement Disorders Clinic and through local support groups to participate in this study. Participants who met the following criteria were included in this study: (1) medical diagnosis of PD [16] (Hoehn & Yahr I-III), (2) self-reported ability to walk >10 m overground without physical assistance, and (3) self-reported ability to walk on a treadmill for 14 minutes (with rest breaks as needed). Potential participants were excluded due to a Hoehn & Yahr Stage IV or V, the presence of uncontrolled cardiorespiratory or metabolic disease (i.e. uncontrolled hypertension, uncontrolled diabetes mellitus), other neurological or orthopedic disorders that may affect walking, or severe communication or comprehension impairments that would impede the ability to perform study procedures appropriately. All participants signed an informed consent form approved by the Institutional Review Board of UNC-Chapel Hill prior to participation. The project was listed on ClinicalTrials.gov (NCT03253965).

Each participant completed the Montreal Cognitive Assessment (MoCA) at the beginning of the testing session to characterize cognitive deficits. They then walked over a 6.1 m (20 foot) Zeno pressure mat (Prokinetics, Haverton, PA) to measure comfortable overground gait speed and cadence without the presence of auditory cues. We used the overground gait speed to guide treadmill speed selection, and the overground cadence to determine the three target frequencies for auditory cues: 85%, 100%, and 115%.

Testing consisted of having all subjects walk during each of the following seven conditions on an instrumented treadmill: (1) no auditory cues (Control), (2) timing their steps to a metronome set to 85% of over-ground cadence, (3) Metronome at 100%, (4) Metronome at 115%, (5) Music at 85%, (6) Music at 100%, and (7) Music at 115%. Each subject selected their own songs that matched the target frequencies from www.bpmdatabase.com. Importantly, participants selected a different song for each Music condition to correspond with the target frequencies. This was necessary to play each song at its correct tempo, rather than artificially slowing down or speeding up a single song. Conditions were block randomized for each participant, by type of auditory cue (music or metronome). Frequencies were randomized within each block. We always conducted the condition with no auditory stimuli (Control) first. Participants were verbally instructed to “step to the beat” of the music or metronome, [17] but no demonstration was provided. With each new condition, participants took ∼15 seconds to adjust to the tempo before we sampled one minute of data. Rest breaks were provided between conditions, as needed. All participants had been on a treadmill at some point previously, but for safety purposes, each subject wore a harness that attached to the ceiling over the treadmill. The harness did not provide any body weight support and did not restrict limb motion. Handrails were available on the sides of the treadmill, though participants were discouraged from using them.

Prior to walking on the treadmill (Bertec Corp, Columbus, OH), a single 14 mm retro-reflective marker was taped to the posterior heel of each shoe. While walking, an eight camera motion capture system (Vicon MX, Los Angeles, CA) sampled the 3D location of both markers at 120 Hz. Ground reaction forces were measured simultaneously using the treadmill’s force plates at 1200 Hz.

### Data Processing & Outcome Measures

Marker trajectories and ground reaction forces were smoothed with a dual-pass 6 Hz and 20 Hz low pass Butterworth filter, respectively. A custom Labview program (National Instruments, Austin, TX) calculated cadence and step length for each condition. Cadence was measured as the inverse of step time, determined from successive heel strikes when the vertical ground reaction force exceeded 20 N. We also computed a cadence ‘error’ as the difference between the measured cadence and the target (intended) cadence. Step length was calculated by measuring the anterior-posterior distance between the right and left heel markers at each heel strike.

### Data Analysis

Data analysis was performed using SPSS (ver 25, IBM, Chicago, IL). We first assessed the cadence error using a two-way repeated measures ANOVA, repeated for cue type (music/metronome) and frequency (85, 100, 115% of overground cadence). The actual cadence was then compared to the intended cadence using paired samples t-tests. We then used a one-way repeated measures ANOVA to assess the specific effect of cueing on cadence compared to the uncued (control) condition. Finally, step length changes were assessed using a two-way repeated measures ANOVA. In the presence of significant main effects, we performed one-way ANOVAs or paired sample t-tests with a Bonferroni correction for post-hoc analyses, as appropriate. Prior to recruitment, we conducted a formal power analysis using G*Power to determine the proposed sample size. Our estimates were based on current literature and expected findings. In particular, Wittwer and colleagues [12] evaluated the effect of both music and a metronome on similar measures in unimpaired older adults. Although the population was different and they did not evaluate different frequencies, we used their results to estimate that we would need 18 participants to observe an effect size of 0.4 with an alpha of 0.05 and power of .80. Given the differences in test populations and the additional measures of frequency, we chose to be conservative and slightly over-recruit.

## Results

Twenty-one individuals with PD participated in this study (13M/8F; age: 69.8±9.8 years, MOCA: 27.0±2.9, UPDRS: 19.4±13.7). With an average time since diagnosis of 8.8±8.1 years, we recruited five individuals classified as H&Y stage 1, nine individuals at stage 2, and seven at stage 3. The average over-ground gait speed was 1.15±0.21 m·s^-1^ and the average treadmill speed used for testing was 1.10±0.26 m·s^-1^ (*p*=.107; d=.37). Except for three participants who walked slower on the treadmill than their typical overground speed, all other participants were able to match their typical overground speed during treadmill testing.

We sought to determine how well participants were able to match their actual cadence with the intended cadence. We observed a cue x frequency interaction in cadence accuracy (*p*=.007, 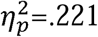, Figure 1). In particular, we noted greater cadence errors produced with music compared to the metronome at the 115% target only (*p*=.003; d=.73). The metronome and music cues produced comparable cadence accuracy at all other target frequencies (*p*>.243). Nevertheless, participants were not able to achieve the intended cadence consistently. Specifically, we observed a higher actual cadence than what was intended for the 85% Metronome (*p*<.001; d=1.07) and 85% Music (*p*<.001; d=1.29) conditions. Additionally, we observed a lower cadence than intended in the 115% Music condition (*p*=.002; d=.75).

**Figure 1:**
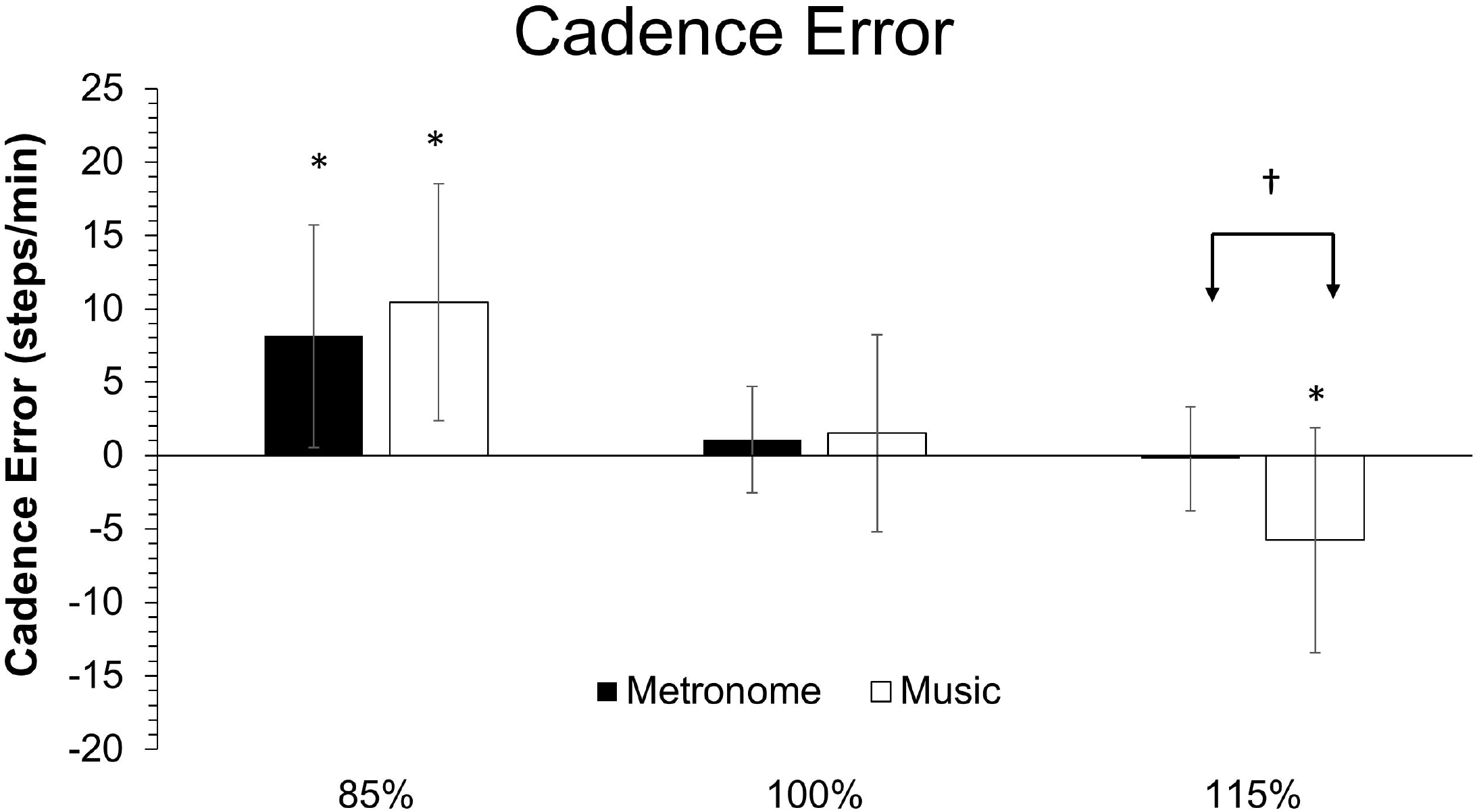
Cadence error is depicted for the metronome (black) and music (white) conditions across frequencies. * = different than intended cadence; † = different between cue conditions. Error bars indicate standard deviations.

Although participants were not consistently achieving the target cadence, the auditory cues influenced cadence compared to walking without auditory cueing (*p*<.001, 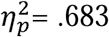, Figure 2). Compared to walking without auditory cueing, participants decreased their cadence while stepping to the 85% Metronome (*p*<.001; d=1.21) and 85% Music (*p*<.001; d=.96) conditions, whereas the 115% metronome condition elicited an increase in cadence (*p*=.004; d=1.47). Cues at 100% of overground cadence (metronome: *p*=1.00; d=.19; music: *p*=1.00; d=.14) and 115% music (*p*=.069; d=.86) did not alter cadence compared to walking without auditory cueing.

**Figure 2:**
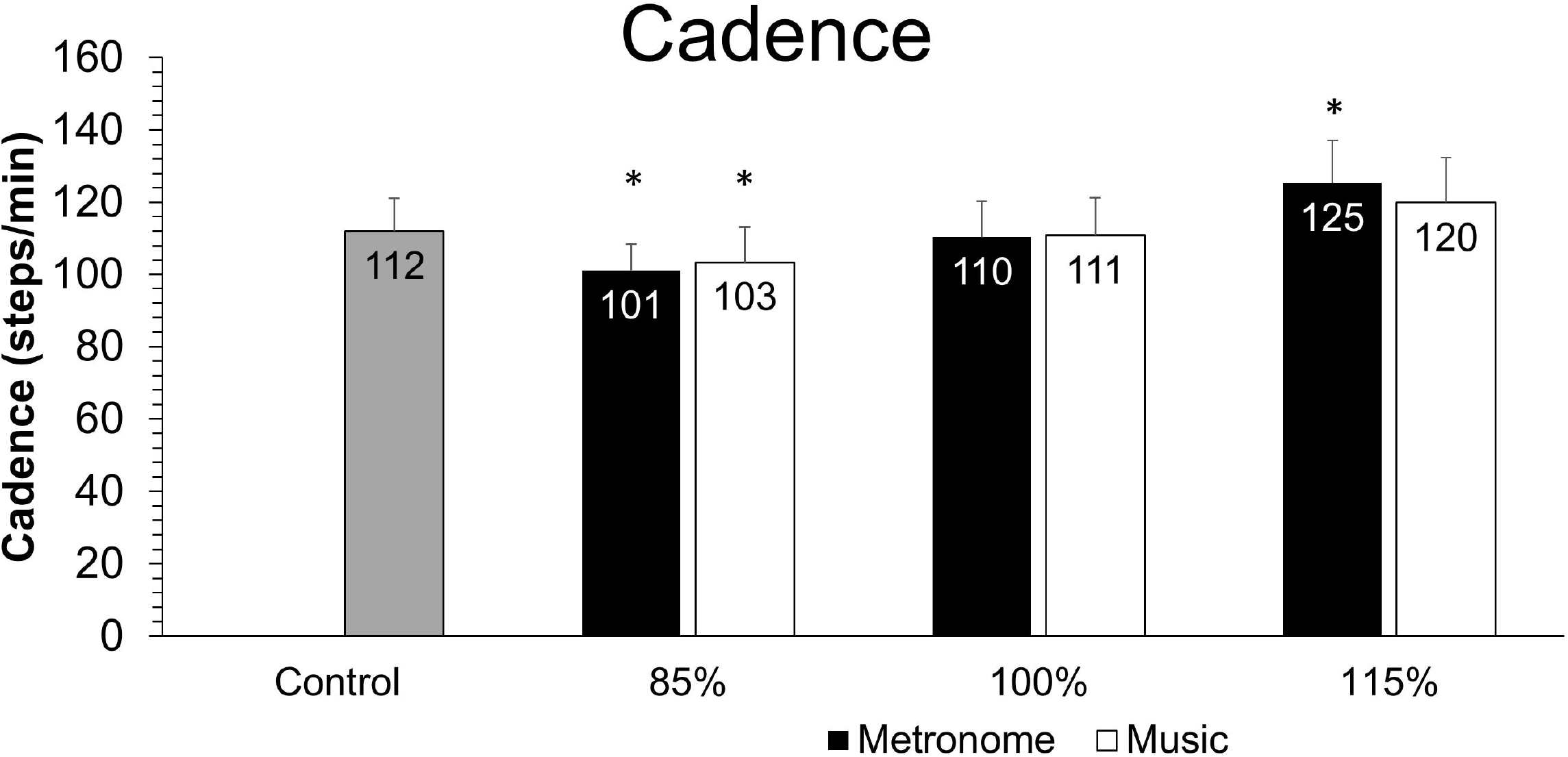
Cadence during treadmill walking without auditory cues (Control, gray bar), and with metronome (black) and music (white) cues. * = different than Control. Error bars indicate standard deviations.

With regards to step length, a cue x frequency interaction was observed (p=.008, 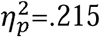, Figure 3). Although there were no differences in step length between the music and metronome at the 85% (*p*=.141; d=.33) and 100% (*p*=.478; d=.16) conditions, we observed that shorter step lengths were observed with the metronome compared to walking with music at the 115% condition (*p*=.004; d=.71). Furthermore, walking with the metronome elicited longer steps during the 85% condition (10% increase; *p*<.001; d=.48), and shorter steps during the 115% condition (11% decrease; *p*<.001; d=.49) compared to walking with no cueing. Likewise, our analysis of walking with music elicited 7% longer steps during the 85% condition (*p*=.003; d=.34), and 6% shorter steps during the 115% condition (*p*=.017; d=.30) compared to walking with no cueing. To put the change into context, we sought minimal detectable change (MDC) values, but were only able to find within-session values for individuals post-stroke [18]. Notably, 19 of the 21 participants exceeded that MDC (i.e., 2.6 cm step length) during one of the 85% conditions, with an average step length increase of 5.7 cm for the 85% metronome condition and 4.0 cm for the 85% music condition.

**Figure 3:**
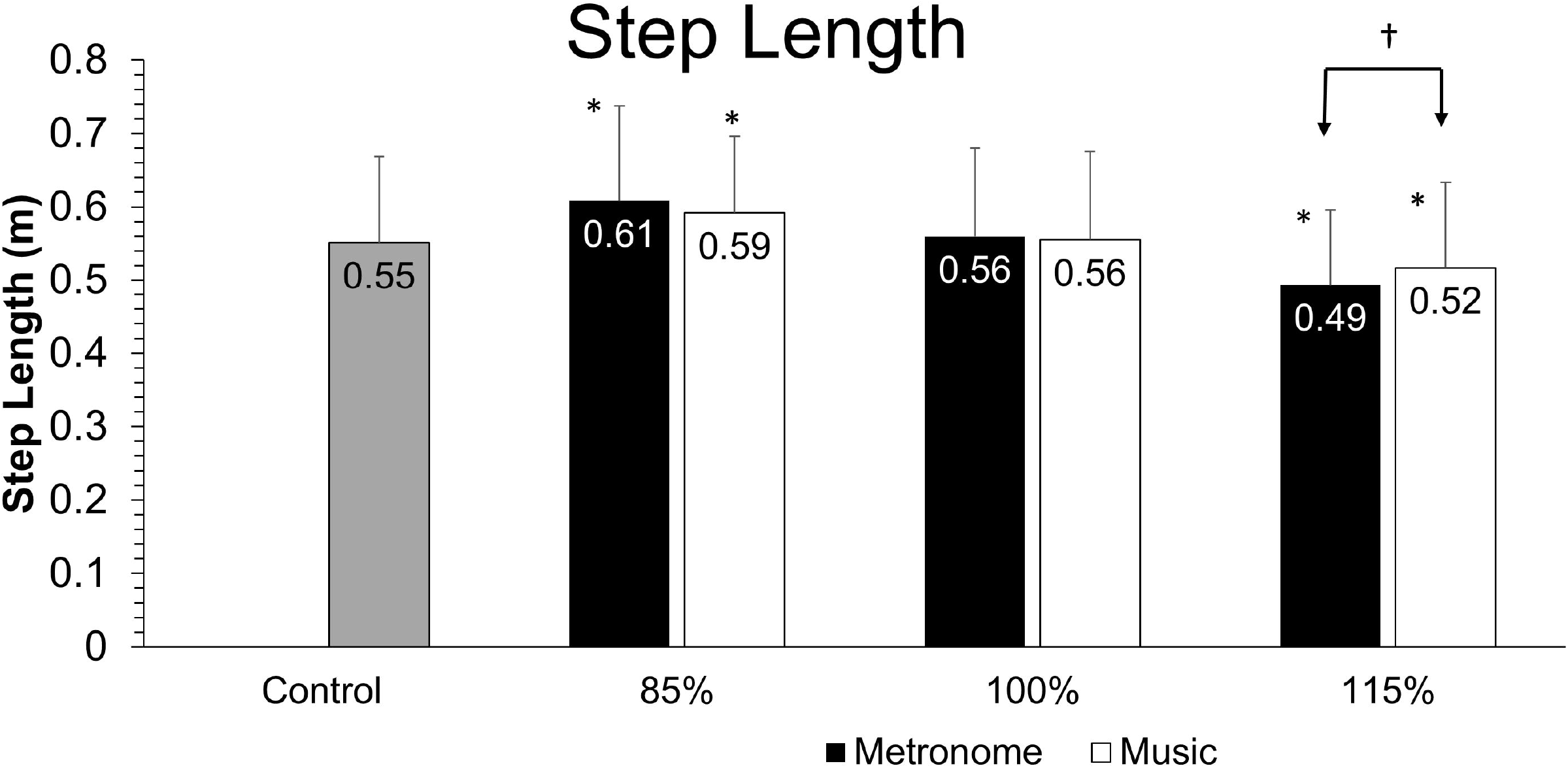
Step lengths during treadmill walking without auditory cues (Control, gray bar), and with metronome (black) and music (white) cues. * = different than Control. † = different between cue conditions. Error bars indicate standard deviations.

## Discussion

This study sought to determine how metronome and music cues of different frequencies would alter spatiotemporal gait parameters for individuals with PD. We observed clear responses based on cueing frequency, but did not observe substantial differences between the metronome and music cues. Therefore, we can reject our hypothesis that metronome cues would elicit a greater response than music cues. We likewise reject our hypothesis that walking with slower frequency cues would not yield a large decrease in cadence. Instead, we found that these slow frequency cues (i.e., 85% of typical overground cadence) produced significantly slower cadence, with the necessary concomitant increase in step length, despite the fact that participants did not match the intended cadence. These findings have implications for identifying parameters of auditory cues that are effective for facilitating longer step length in individuals with PD during treadmill ambulation.

Step lengths are already notably shorter in individuals with PD [19]. Thus, therapists require approaches that have the capability of reliably increasing step length. Indeed, we found that walking on a treadmill with a slower (i.e., 85%) cue frequency generates a longer step length, whereas faster cues (115%) lead to a shorter step length. Our findings may appear to contrast with previous studies that demonstrated that *faster* cadence cues yield increased stride length,[20] or that stride length does not change with cue frequency [21-23]. Of note, however, is that these prior studies were performed overground and the resulting ability to change gait speed could contribute to these discrepant findings. The results of our study extend prior work by demonstrating that when we control speed with the treadmill, people with PD are able to take consistently longer steps with exposure to metronome cues at frequencies *slower* than their comfortable cadence. Future work will be needed to generalize these results to an overground environment.

Prior work on auditory cueing advocates for the use of higher frequencies to increase stride length and gait speed [5, 10, 24]. However, our data clearly suggest that if individuals with PD practice walking on a treadmill, the use of higher frequency cues will result in *shorter* steps, which may exacerbate their deficits. This is particularly relevant, because therapists commonly use treadmills to provide relevant external physical cues to enhance the automaticity of gait [25] and promote consistent stepping practice [26, 27]. Coupling treadmill walking with higher frequency cues, however, would be counterproductive. Instead, we propose that pairing slower auditory cues with treadmill walking would encourage longer steps.

For the purposes of applying these findings to therapy for individuals with PD, it is noteworthy that we observed increases in step length, despite participants’ inability to synchronize their steps to the beat. Surprisingly, the inability to step at the intended frequency occurred with both music and metronome cues, and suggests that the observed error may have had little to do with the cue itself. Instead, we attribute the cadence error to participants underlying timing impairment [3, 28]. Research involving unimpaired participants suggests that the ability to synchronize steps to rhythmic auditory cues is improved with faster music and music that the individual is familiar with [7, 29]. In our cohort of participants with PD, we can speculate that the increased cadence error at slower frequencies was due to the difficulty in concurrently having to take a longer step. Such deficits in step length regulation may be due to changes in the ‘gain’ of motor function [14], deficits in motivation or reward circuitry in the basal ganglia, or a speed/accuracy tradeoff that influences energy cost [30].

Previously, musical cues have been associated with improvements in gait speed, cadence, and stride length [9, 10, 24]. It was suggested that the continuity of sound inherent in musical cues requires less cognitive demand than listening to cues that have a fixed beat, such as a metronome [5]. Nevertheless, participants subjectively reported greater difficulty identifying the tempo during music cues compared to the metronome cues. It is possible that some of the songs were considered ‘low-groove music’ or that some of our participants exhibited deficits in beat perception, which we did not measure [7]. Despite their perceived difficulty in identifying the musical beats, we failed to identify consistent differences in gait parameters when using the metronome or music cues. These data suggest that music may serve as a suitable surrogate to a metronome in clinical practice.

Importantly, prior studies often used musical tracks that were designed specifically to highlight the beat of the music to enable easy detection of the intended rhythm [11, 24]. This is not always feasible or enjoyable to implement in the clinical or community setting. For this reason, we allowed participants to select their own songs that coincided with the intended frequency. Familiar music creates greater entrainment than unfamiliar music, perhaps due to less cognitive demand [29]. Further, by allowing participants to select their own music, we believe that we have a better representation of the effect of music use in a real world setting. Because many people like to listen to music when they exercise or are active, this pragmatic approach is easily translatable to clinical settings.

### Limitations

There were several limitations to this study. First, is that some participants held onto the rails while completing the walking trials on the treadmill. Although handrail use may alter stride lengths for people with PD [26], all individuals requiring handrails used them consistently across all conditions. This should presumably limit the effect on the within-subject comparisons that we performed. Secondly, each participant selected different songs from potentially different genres. It is also possible that we started these songs in a section where the beat may have been difficult to distinguish. Because of our pragmatic design, we were unable to control this limitation adequately without selecting a single song for use across subjects and conditions. Finally, we tested a different number of participants for each Hoehn and Yahr stage, resulting in a slightly larger number of participants classified as Stage 2 or 3. As a result, our results may not generalize as well to individuals classified as Stage 1.

In conclusion, our study demonstrated that walking on a treadmill at constant speed with *slower* auditory cueing could elicit reductions in cadence (with concomitant longer step lengths) in patients with PD. This important change conflicts with conventional approaches, and was apparent despite participant’s inability to synchronize their steps fully to the auditory cues. The music and metronome cues produced comparable changes to gait, suggesting that either form of cue may be effective at overcoming the shortened step lengths that are prevalent for individuals with PD.

## Data Availability

Data will be made available upon request.

## Acknowledgment

This research did not receive any specific grant from funding agencies in the public, commercial, or not-for-profit sectors.

## References

1. Morris ME, Martin CL, Schenkman ML. Striding out with Parkinson disease: evidence-based physical therapy for gait disorders. Physical therapy 2010; 90: 280–8.

2. Spaulding SJ, Barber B, Colby M, Cormack B, Mick T, Jenkins ME. Cueing and gait improvement among people with Parkinson’s disease: a meta-analysis. Archives of physical medicine and rehabilitation 2013; 94: 562–70.

3. Wu T, Hallett M, Chan P. Motor automaticity in Parkinson’s disease. Neurobiology of disease 2015; 82: 226–234.

4. Keus SH, Munneke M, Nijkrake MJ, Kwakkel G, Bloem BR. Physical therapy in Parkinson’s disease: evolution and future challenges. Mov Disord 2009; 24: 1–14.

5. Ashoori A, Eagleman DM, Jankovic J. Effects of Auditory Rhythm and Music on Gait Disturbances in Parkinson’s Disease. Frontiers in neurology 2015; 6: 234.

6. Willems AM, Nieuwboer A, Chavret F, Desloovere K, Dom R, Rochester L, et al. The use of rhythmic auditory cues to influence gait in patients with Parkinson’s disease, the differential effect for freezers and non-freezers, an explorative study. Disabil Rehabil 2006; 28: 721–8.

7. Leow LA, Parrott T, Grahn JA. Individual differences in beat perception affect gait responses to low- and high-groove music. Frontiers in human neuroscience 2014; 8: 811.

8. Rodger MW, Craig CM. Beyond the Metronome: Auditory Events and Music May Afford More than Just Interval Durations as Gait Cues in Parkinson’s Disease. Frontiers in neuroscience 2016; 10: 272.

9. Bella SD, Benoit CE, Farrugia N, Keller PE, Obrig H, Mainka S, et al. Gait improvement via rhythmic stimulation in Parkinson’s disease is linked to rhythmic skills. Scientific reports 2017; 7: 42005.

10. Ford MP, Malone LA, Nyikos I, Yelisetty R, Bickel CS. Gait training with progressive external auditory cueing in persons with Parkinson’s disease. Archives of physical medicine and rehabilitation 2010; 91: 1255–61.

11. McIntosh GC, Brown SH, Rice RR, Thaut MH. Rhythmic auditory-motor facilitation of gait patterns in patients with Parkinson’s disease. J Neurol Neurosurg Psychiatry 1997; 62: 22–6.

12. Wittwer JE, Webster KE, Hill K. Music and metronome cues produce different effects on gait spatiotemporal measures but not gait variability in healthy older adults. Gait & posture 2013; 37: 219–22.

13. Lewek MD. The influence of body weight support on ankle mechanics during treadmill walking. J Biomech 2011; 44: 128–33.

14. Morris ME, Iansek R, Matyas TA, Summers JJ. The pathogenesis of gait hypokinesia in Parkinson’s disease. Brain 1994; 117 (Pt 5): 1169–81.

15. Morris ME, Huxham F, McGinley J, Dodd K, Iansek R. The biomechanics and motor control of gait in Parkinson disease. Clin Biomech (Bristol, Avon) 2001; 16: 459–70.

16. Postuma RB, Berg D, Stern M, Poewe W, Olanow CW, Oertel W, et al. MDS clinical diagnostic criteria for Parkinson’s disease. Mov Disord 2015; 30: 1591–601.

17. Mendonca C, Oliveira M, Fontes L, Santos J. The effect of instruction to synchronize over step frequency while walking with auditory cues on a treadmill. Human movement science 2014; 33: 33–42.

18. Kesar TM, Binder-Macleod SA, Hicks GE, Reisman DS. Minimal detectable change for gait variables collected during treadmill walking in individuals post-stroke. Gait & posture 2011; 33: 314–7.

19. Morris ME, Iansek R, Matyas TA, Summers JJ. Stride length regulation in Parkinson’s disease. Normalization strategies and underlying mechanisms. Brain 1996; 119 (Pt 2): 551–68.

20. Picelli A, Camin M, Tinazzi M, Vangelista A, Cosentino A, Fiaschi A, et al. Three-dimensional motion analysis of the effects of auditory cueing on gait pattern in patients with Parkinson’s disease: a preliminary investigation. Neurological sciences : official journal of the Italian Neurological Society and of the Italian Society of Clinical Neurophysiology 2010; 31: 423–30.

21. Howe TE, Lovgreen B, Cody FW, Ashton VJ, Oldham JA. Auditory cues can modify the gait of persons with early-stage Parkinson’s disease: a method for enhancing parkinsonian walking performance? Clin Rehabil 2003; 17: 363–7.

22. Suteerawattananon M, Morris GS, Etnyre BR, Jankovic J, Protas EJ. Effects of visual and auditory cues on gait in individuals with Parkinson’s disease. J Neurol Sci 2004; 219: 63–9.

23. Lohnes CA, Earhart GM. The impact of attentional, auditory, and combined cues on walking during single and cognitive dual tasks in Parkinson disease. Gait & posture 2011; 33: 478–83.

24. Thaut MH, McIntosh GC, Rice RR, Miller RA, Rathbun J, Brault JM. Rhythmic auditory stimulation in gait training for Parkinson’s disease patients. Mov Disord 1996; 11: 193–200.

25. Frazzitta G, Maestri R, Uccellini D, Bertotti G, Abelli P. Rehabilitation treatment of gait in patients with Parkinson’s disease with freezing: a comparison between two physical therapy protocols using visual and auditory cues with or without treadmill training. Mov Disord 2009; 24: 1139–43.

26. Frenkel-Toledo S, Giladi N, Peretz C, Herman T, Gruendlinger L, Hausdorff JM. Treadmill walking as an external pacemaker to improve gait rhythm and stability in Parkinson’s disease. Mov Disord 2005; 20: 1109–14.

27. Bello O, Marquez G, Camblor M, Fernandez-Del-Olmo M. Mechanisms involved in treadmill walking improvements in Parkinson’s disease. Gait & posture 2010; 32: 118–23.

28. Baltadjieva R, Giladi N, Gruendlinger L, Peretz C, Hausdorff JM. Marked alterations in the gait timing and rhythmicity of patients with de novo Parkinson’s disease. Eur J Neurosci 2006; 24: 1815–20.

29. Leow LA, Rinchon C, Grahn J. Familiarity with music increases walking speed in rhythmic auditory cuing. Ann N Y Acad Sci 2015; 1337: 53–61.

30. Mazzoni P, Hristova A, Krakauer JW. Why don’t we move faster? Parkinson’s disease, movement vigor, and implicit motivation. J Neurosci 2007; 27: 7105–16.

